# A potential transition from a concentrated to a generalized HIV epidemic: the case of Madagascar

**DOI:** 10.1101/2022.09.26.22280390

**Authors:** David Alonso, Xavier Vallès

## Abstract

HIV expansion is controlled by a range of interrelated factors, including the natural history of HIV infection and socio-economical and structural factors. However, how they dynamically interact in particular contexts to drive a transition from concentrated HIV epidemics in vulnerable groups to generalized epidemics is poorly understood. We developed a compartmental dynamic model using available data from Madagascar, a country with a contrasting concentrated epidemic, to explore the interaction between these factors with special consideration of commercial and transactional sex as HIV-infection drivers. The model predicts sigmoidal-like prevalence curves with turning points within years 2020-2022, and prevalence reaching stabilization by 2033 within 9 to 24% in the studied (10 out of 11) cities, similar to high-prevalence regions in Southern Africa. Circumcision could have slowed down HIV propagation, but, given the key interplay between risky behaviors associated to young women and acute infections prevalence, mediated by transactional sex, the protective effect of circumcision is currently insufficient to contain the expansion of the disease in Madagascar. These results suggest that Madagascar may be experiencing a silent transition from a concentrated to a generalized HIV epidemic. This case-study model could help to understand how this HIV epidemic transition occurs.

## Introduction

In 1981 the first cases of AIDS were described in San Francisco (USA), and the infectious nature of the new entity was soon established. By 1984 the causal pathogen, the HIV, was characterized, and the transmission mechanisms were described. By then, the world realized that the infection was already widespread in Sub-Saharan Africa, where the majority of cases still occur and, more specifically, in Southern-Africa with a pattern of generalized epidemics. However, reasons why some countries or regions in Sub-Saharan Africa show disparate epidemic profiles remain unclear. Although the mechanisms that drive HIV expansion are well-established, how they interact to shape the transition from low to high prevalence and sustained HIV incidence has not been fully elucidated, but has important implications for guiding responses [1]. Concentrated HIV epidemic are defined by the occurrence of the infection largely in identified vulnerable groups, such as sex workers, men who have sex with men, and injected drugs users. Conversely, HIV epidemic is termed as generalized when the transmission is sustained in the general population, defined as a general population prevalence of over 1% or in a sentinel population like pregnant women. The transition from concentrated to generalized epidemic would take place if R0 is over 1 for a long period of time with self-sustained transmission within the general population. From a public health perspective, once the HIV epidemic has a generalized profile, it would likely persist despite effective programs focusing on vulnerable groups.

In this study, we develop a new mathematical model to explore the mechanisms driving such transition, using Madagascar as a case-study. The island of Madagascar (formally Republic of Madagascar) is located in the east coast of Africa, close to Mozambique and South-Africa. By contrast to Southern Africa, Madagascar shows a low prevalence of HIV in the general population (less than 1%), alongside a high HIV prevalence among key populations. This is an astonishing concentrated epidemic profile, particularly given that Madagascar shows a widespread presence of the most recognized risk factors associated with HIV acquisition [2]. Since the late 90’s, previous studies have predicted that Madagascar was near the tipping point towards a generalized epidemic [2, 3]. The transition has not occurred to date, even if the general trend is an increase of prevalence among key populations [2].

## Methods

### The HIV Transmission Model

We represent the temporal dynamics of disease spread by a set of ordinary differential equations [4]. This system represents the progression of the disease as a consequence of sexual encounters between infectious and non-infected individuals (see Fig 1).

**Figure 1:**
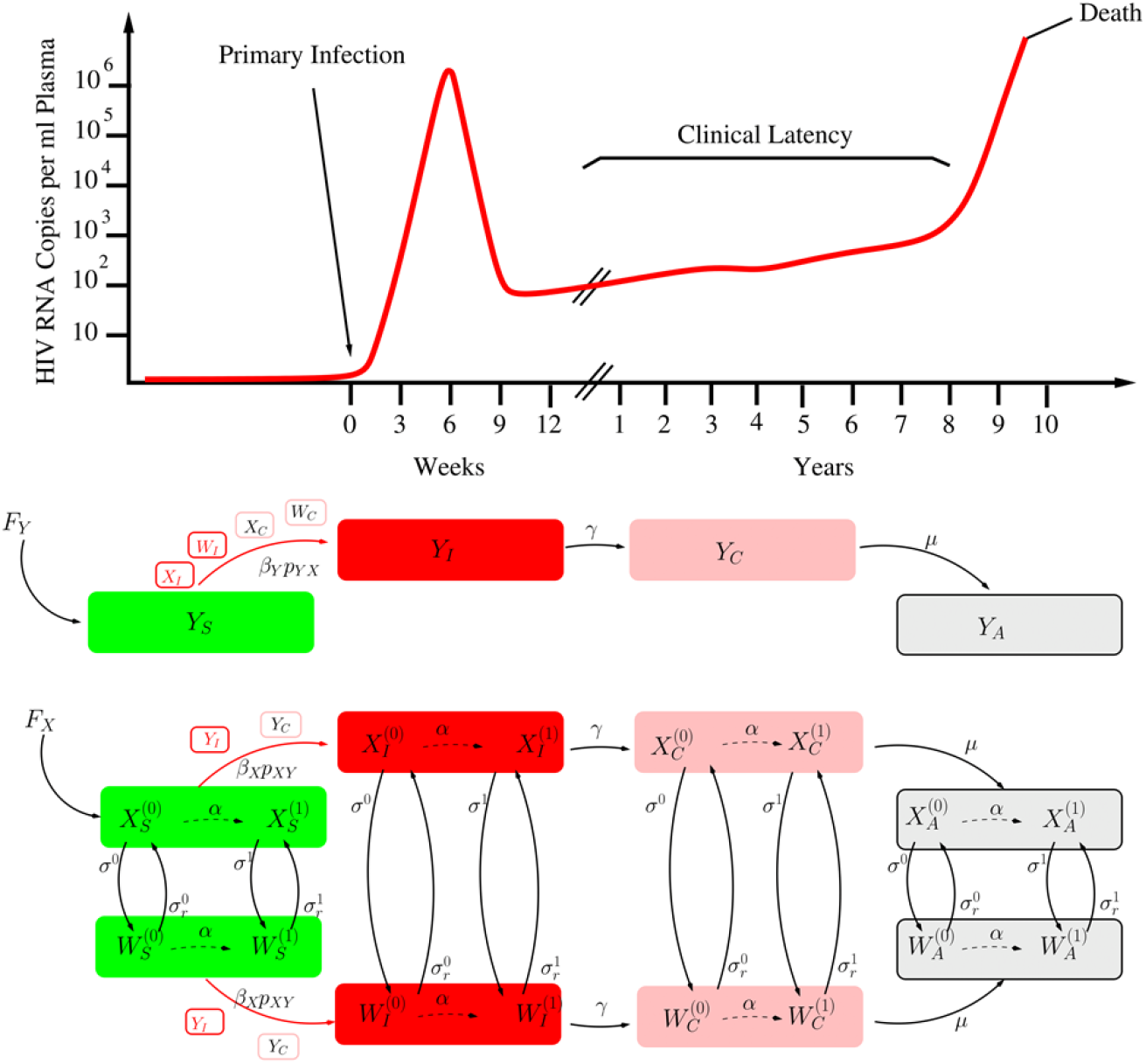
Graphic representation of women and men subpopulations progressing through the different stages of the disease since they acquire the infection from infectious males and women, respectively^1^ ^1^At any stage of the disease, women (X) can become sexual workers (W), at rate σ, or reverse that condition, at rate σr. The aging rate α controls the transition from young (see superscript (0)) to adult women (see superscript (1)). All stages, both in men and women, are subject to mortality. Additional disease-induced mortality is only considered during the AIDS phase (see subscript A). For simplicity, arrows representing this fatal transition are not shown. The typical course of a HIV infection is drawn on the top panel.

The whole population is divided into a set of groups. The male population is considered as a single group, while female population is subdivided into four groups: two groups according to sexual activity (sexual workers and rest of women), where each of them is in turn subdivided into young Adolescent Girls and Young Women or AGYW (women between 15 to 24 years old) and adult females. The particular consideration of AGYW is based on the recognition of their high vulnerability to HIV in Sub-Saharan Africa (see discussion) [5]. Both males and females are recruited into the population as fully susceptible individuals, represented by the subscript S in the set of dynamic variables [see Eqs (SA1)-(SA11) in SI.]. These recruitment rates are defined as the number of males and females per unit time that reach sexual maturity at any given time. As these individuals encounter infectious sexual partners, they can acquire the infection and then transition to the HIV acute infection stage (see subscript (I) in Fig 1 and equations in SI). This stage lasts, on average, between 2 and 3 months [6, 7] (see values in Table SA1 in SI), and is characterized by a transient high viral load and infectiousness (Fig 1) [7, 8]. The following stage (chronic or stable stage) is characterized by a much lower viral loads and infectiousness. The duration (see subscript C) is controlled by the rate μ (see Fig 1), and lasts around 10 years [7, 9] followed by the breakdown of the immune system leading into the AIDS stage (see subscript A). See the supporting information (SI) for a comprehensive description of the system of ODEs.

### The Demographic Model

In the absence of disease transmission, female and male adult subpopulations grow as a result of a balance between a recruitment rate into sexual maturity and an average mortality rate. Adult populations are driven by time-dependent demographic parameters (recruitment rates, FX and FY, and adult mortality rates, dX and dY). FX and FY represent the absolute number of females and males entering sexual active life per year, respectively. They depend on the population size of every city and have a monotone increasing trend over time, which along with immigration from rural areas, leads to demographic growth (see the case of Antananarivo in Fig SC1). In the supporting information, we explain the strategy we followed to accurately estimate them from demographic life table data. The rest of model demographic parameters (α and σ’s, see Fig 1 and Table SA1 in SI) were searched to be compatible with surveyed sex worker population across cities in 2014 and 2017. We fit a trajectory for the number of sex workers that could have been observed from 2000 to 2016 under two reasonable hypotheses, namely, the constant-fraction and the sigmoidal hypotheses. The first one assumes that the average fraction of sex workers in the population remained constant over the whole period of interest (2000-2016).The second hypothesis takes into consideration Madagascar economic crises (2009-2013) and makes the assumption that the fraction of sexual workers within the adult female population could have increased as a consequence of the crisis. We modeled this growth as a sigmoidal curve (see SI for details).

### Model Validation

We used disease data (see Table SF1 in SI) and the demographic information found in annual life tables from 2000 to 2016 to search for parameter combinations able to yield temporal trajectories in agreement with the HIV epidemiological data available. The process of model assessment and validation was done in three different phases: (1) the simple demographic model [see Eqs. (SA13) from the SI], (2) the expanded demographic model [see Eqs. (SA12) from the SI] and, finally (3) the full disease transmission model [see Eqs. (SA1)-(SA11) from the SI]. Parameter estimations were based on several data sources [10–13]. Initial population values were chosen to correspond to the year 2000. Adult sex ratio is considered 1:1 [12]. Disease initial HIV prevalence for sexual workers in 2000 was set to be a 1% of the 2016 prevalence value for each city (see Table SF1 and SF2 in SI) because these yield very low initial numbers of infected individuals in 2000 (between 1 and 10 for most cities). Disease prevalence for the rest of groups at the initial year are then set accordingly. Parameter distributions consistent with both demographic and disease data showed that model parameter values were further constrained by population and disease data over the studied period (see Fig. SE7 in SI).

### Model Projections

The ensemble of parametric configurations providing good fit to data up to 2016 (see Table B1 and B2 in SI) was then used to project the evolution of the disease up to 2033. The numerical integration of the full system requires annual time-dependent parameters, this is, future recruitment and mortality rates from 2016 to 2033. These were extrapolated from the same demographic life tables under the assumption that mortality and fertility rates maintain the trends observed between 2000 and 2016. This procedure yielded expected annual rates (FX, FY, dX, and dY) up to 2033 (see Fig. SE1 in SI). Model projections were then calculated from year 2016 up to year 2033 taking into account the uncertainty we have in model parameter estimates (see SI for details).

### Data availability

Base and intermediate data used for this study is fully available in the referenced link as a Dryad dataset [14].

## Results

### R0 and the Stationary State

The average R0 values across the 10 studied cities are shown in Fig 2 (see also Tables SB1 and SB2 in SI). Since some parameters are time-dependent, these values correspond to year 2000, and are calculated using Eq (SB46) (see SI). They appear to be consistently bounded between 3 and 9 across the different cities. We plot R0 in year 2000 in terms of male sexual encounter rates and the probability (pY X) of acquiring the infection for a male encountering an infectious female for the city of Antananarivo (Fig 3). This allows picturing the effect on R0 of circumcision, which would reduce this infection probability. In no city, the conservative estimation of 60% risk reduction due to circumcision [15–17] of HIV transmission from female to male (pY X) would have taken R0 below 1 (see also Figs. SB1 and SB2 in SI). We also explore the effect of circumcision in Fig 4, where we compare model predictions when we use a constrained prior (0 < pY X < 0.0004) vs using a less constrained one (0 < pY X < 0.001) in our parameter searches (see also Fig SE4 and SE5 in SI). If R0 > 1 and parameters were kept constant, stationary prevalence values would be reached. This stationary state can be calculated as well by using a semi-analytical approach [18] (see Fig SA1 in SI).

**Figure 2:**
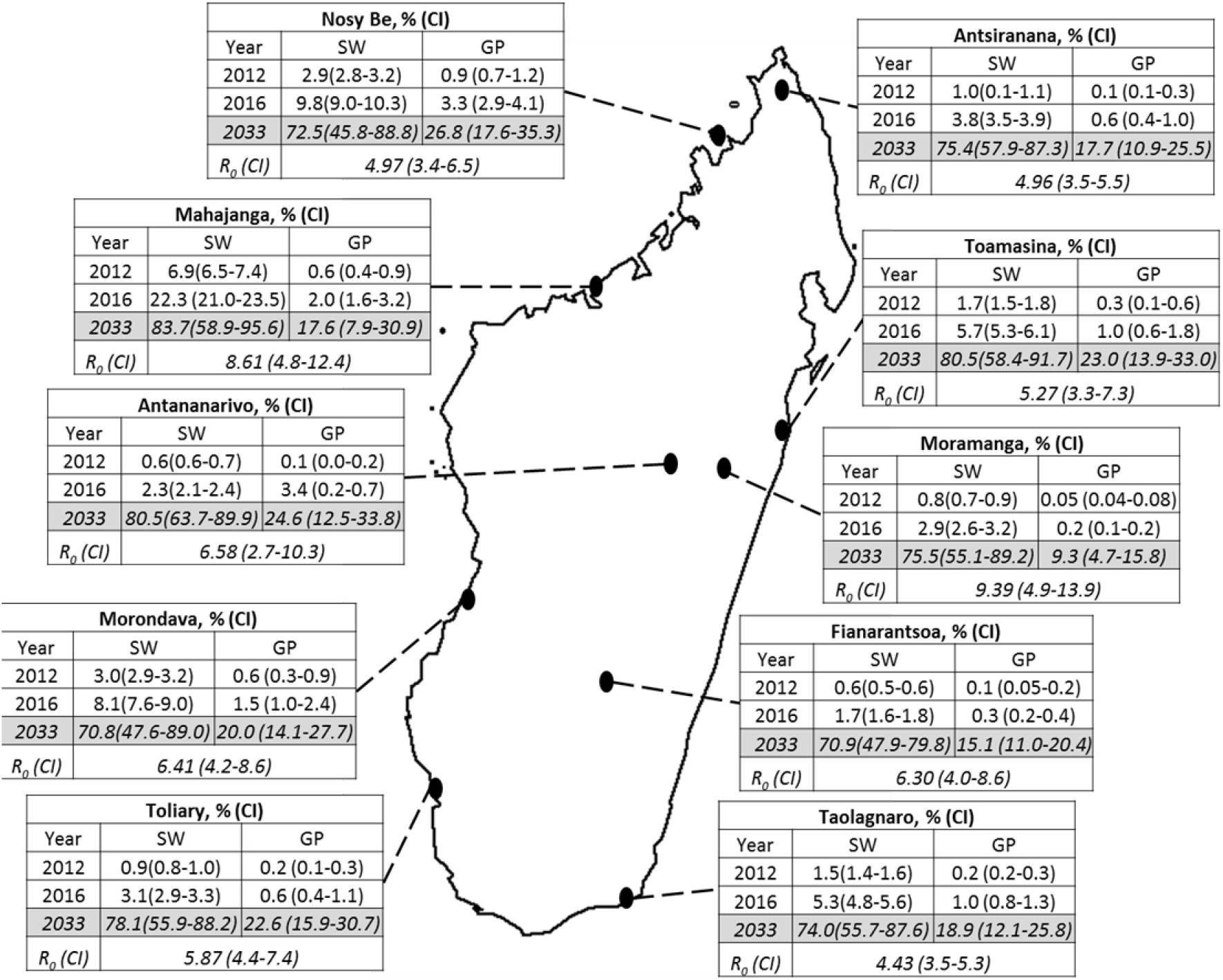
R0 values in 2000 and HIV prevalence levels both in the general (GP) and in the SW population for the two last years for which surveyed data existed (2012 and 2016, see Table SF1 in SI) along with our model prediction for 2033 (see shaded row)^1^. 1. Figure modified from [2] with permission from the authors.

**Figure 3:**
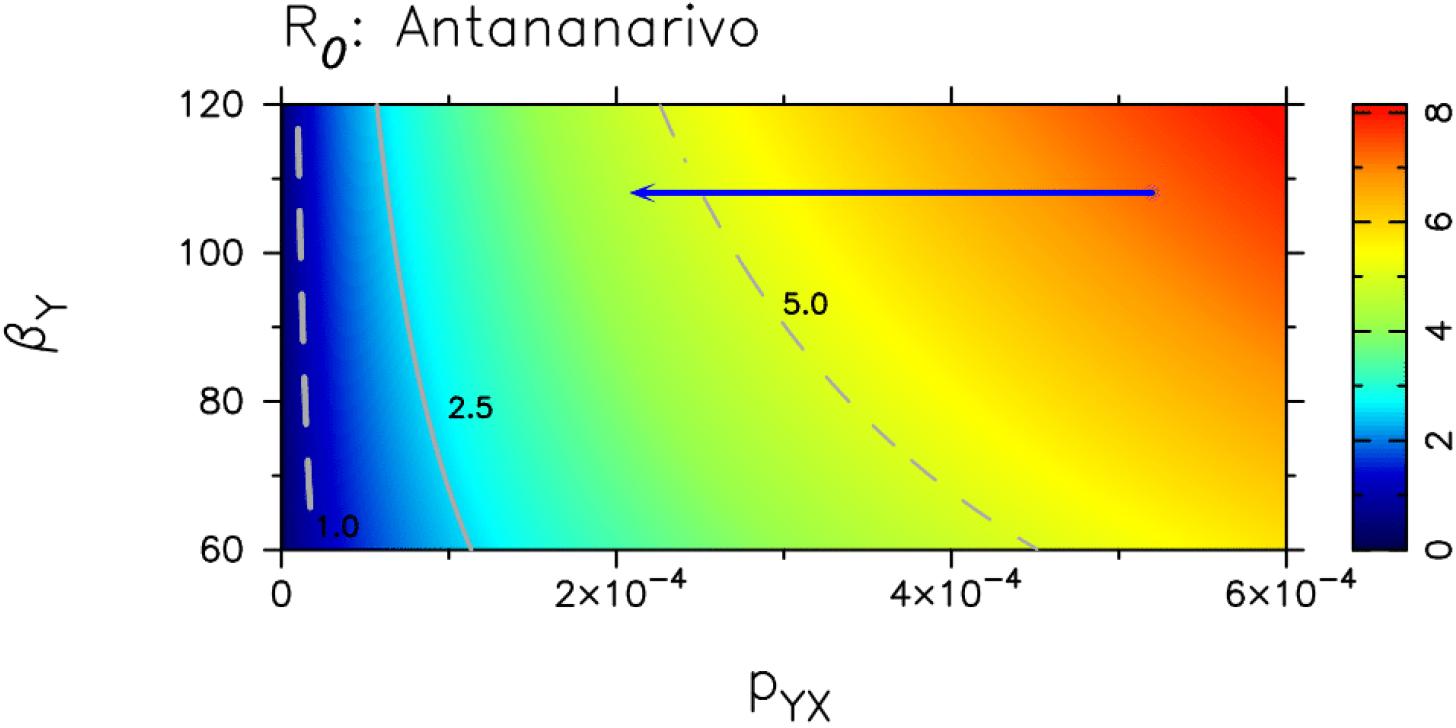
R0 changes as a function of βY, the sexual encounter rate of males, and pY X, the transmission probability from infected females to males^1^. 1. The rest of model parameters, but βY and pY X, are kept constant. Their values are chosen to correspond to Antananarivo (see Table SB1, and also Figs. SB1 and SB2 in SI). Time-dependent parameters FX, FY, dX and dY are set to their values in 2000. The estimated average female-to-male transmission probabilities and the sexual encounter rates (β_Y_) are represented by a little circle defining the initial coordinates of the arrow. The tip of the arrow represents the potential reduction in R0 caused by a 60% reduction in the transmission probability from infectious females to males as a consequence of circumcision (as reported in [14–16]).

**Figure 4a and 4 b:**
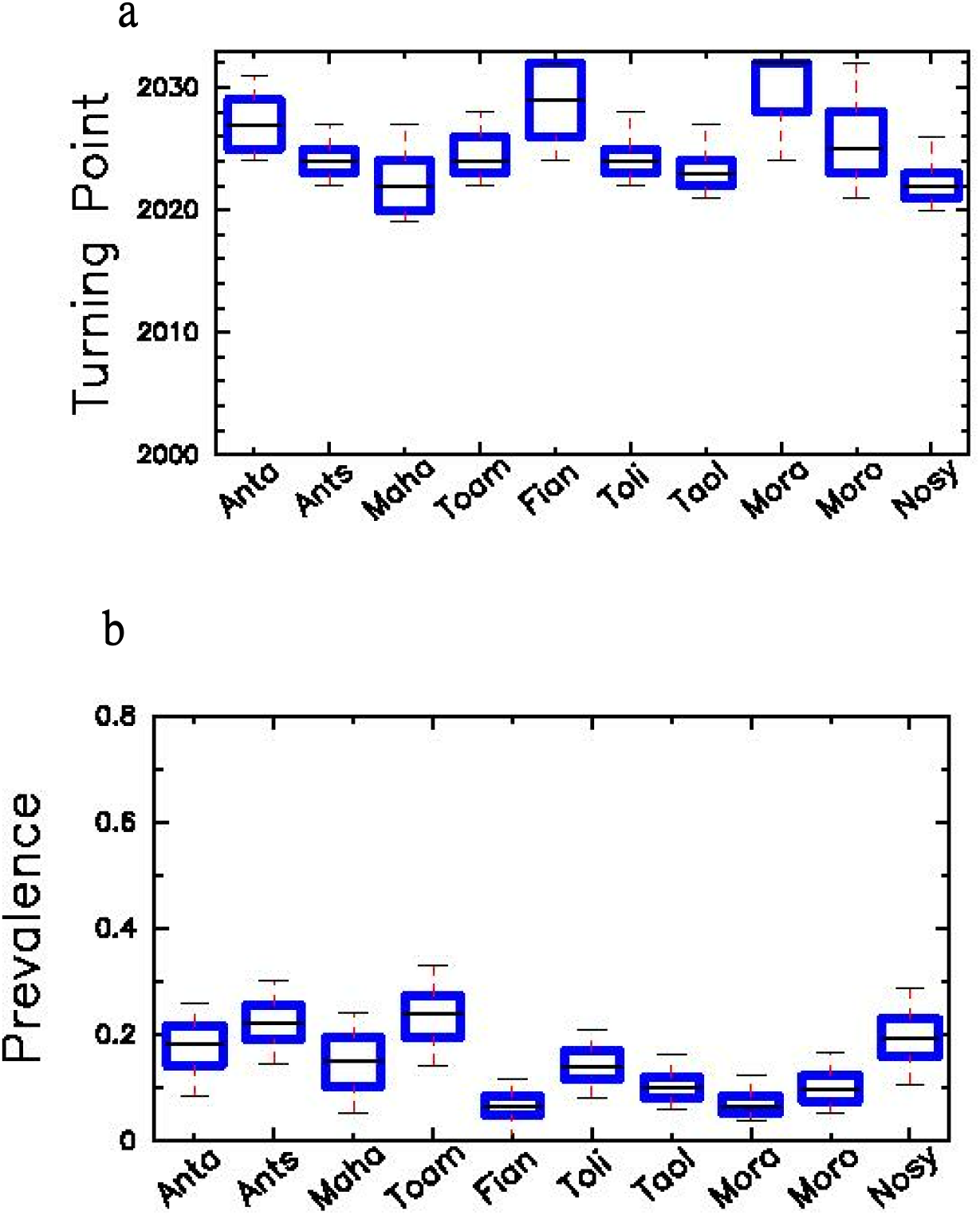
Turning points (blue, left vertical axis) and expected prevalence in 2033 (magenta, right vertical axis) calculated for the projected temporal evolution of total disease prevalence within the general population in the 10 cities^1^. 1. Box plots represent distributions across the parameter configurations that provided a good fit to data for the period 2000-2016. In Fig 4a, parametric configurations were searched within pY X values between 0.0 and 0.001, as indicated in Table SA1 in SI, while, in Fig 4b, searches were conducted by constraining even more pY X (between 0.0 and 0.0004), to mimic the effect of circumcision as a 60% reduction in transmission from females to males.

### Model Projections

The model shows sigmoidal-like dynamics of disease establishment (see Fig 5 and Figs. SE2-SE6 in SI) upon introduction, which can be characterized by three different phases. The introduction or initial phase may last about 15 to 25 years. This period is characterized by a slight exponential increase in prevalence, but still at very low levels. After this initial phase, the disease finally takes off. Full disease establishment is only reached after about 35 to 55 years since the disease was initially introduced. A sharp transition between low to high prevalence characterizes the intermediate or transition phase, which is fast and may only last about 10 years (see Fig SA2 in SI).These overall temporal scales are a direct consequence of the estimated parametric configurations within biologically reasonable ranges (see average values in Tables SB1 and SB2 in SI). To characterize these phases, we defined two thresholds and a turning point (the time at which the rate of increase along the projected temporal evolution is the highest). The first threshold which is the end of the initial phase is set when prevalence is 10% of the maximum value. The second threshold which is the beginning of the third phase is set when prevalence has reached 90% of the corresponding maximum value. Maximum prevalence is defined as the prevalence level reached by the end of the projected period (2033). Turning-point year distributions and the prevalence levels in the whole population for the ensemble of parametric configurations are shown in Fig 4a and 4b.

**Figure 5a and 5b:**
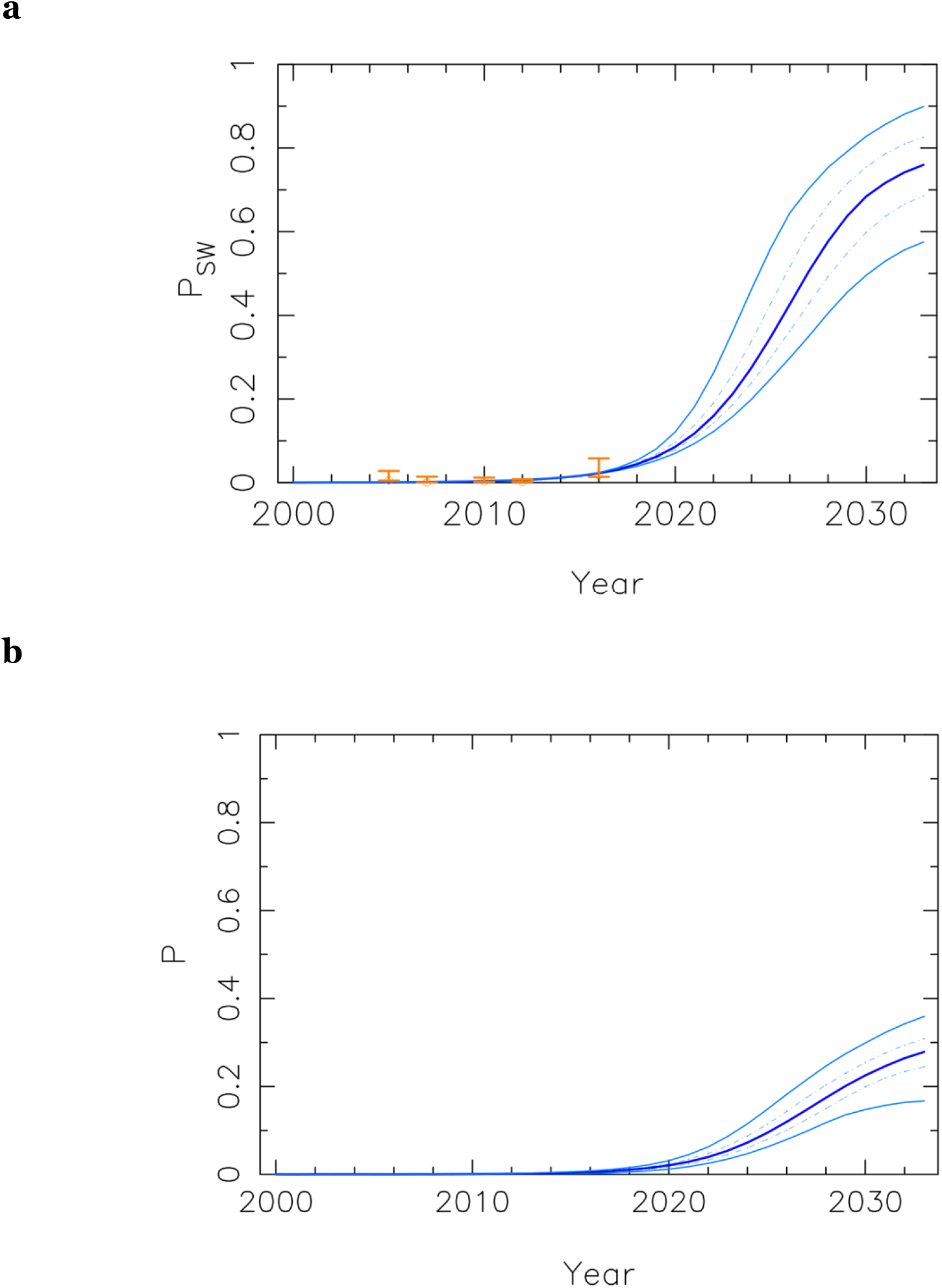
Projected prevalence trends in Antananarivo, as a fraction of infected individuals, for the SW (PSW) and overall adult (P) populations are represented^1^. 1. True observed data (for 2005, 2007, 2010, 2012 and 2016) are highlighted (in orange) in panel (a), with error bars representing confidence intervals. The five lines represent 5%, 25%, 50%, 75%, and 95% percentiles from the lowest to the highest values, respectively.

Disease prevalence for every key group follows a slightly different trajectory from introduction to full establishment, being always the sex workers prevalence the first to take off, and the one reaching the highest values. For comparison, we show the temporal evolution of the prevalence within sex workers and the general population in Antananarivo (Fig 5).

Other cities showed a similar pattern (see Figs. SE2-SE6 in SI). Our model predicts a sharp increase in prevalence values around 2022 until reaching steady values over 2030 (see Figs 4 and 5). This pattern is consistent across cities (see also Figs SE2-SE6 in SI). In Fig 5a, we show, for instance, an increase of disease prevalence within the sex worker key population jumping from average values of 1% before 2015 to values as high as 60% after 2030, while overall prevalence in the total population stabilize around values from 20% to 30% (Fig 5b). We recall here that overall prevalence is given as a fraction of total adult population. Also, we visually show a summary of projected prevalence in 2033 in the general population along with of R0 values in 2000 across cities over the map of Madagascar (see Fig 2).

## Discussion

Our model supports the notion that a complex interplay between different drivers determines the timing and speed of the transition from a concentrated to a generalized HIV epidemic across different countries and sub-regions. In Madagascar, we believe that two major factors may have been modulating disease expansion, and delaying this potential transition: the late/slow introduction of HIV and/or widespread practice of circumcision. According to our analyses, HIV prevalence should have been very low still in 2000 according to the earliest data available from 2005 [19] and 2007 [20], but estimated R0 was already much larger than 1 in most cities. Therefore, the model reveals that the protective effect of circumcision was minimal back then, and may have already vanished in Madagascar (Fig 3). This is in agreement with previous authors, which pointed out that over certain threshold of HIV prevalence among sex workers, circumcision does not have a substantial effect towards HIV prevention at population level [21]. Furthermore, our results suggest that the turning point towards generalized epidemic in Madagascar is very close or may have even been surpassed in certain localities (Fig4). At this stage, AGYW may play a key role mediated by transactional sex in the generalization and maintenance of the epidemics in the general population (Fig 6), with a key interaction with acute HIV infections.

**Figure 6:**
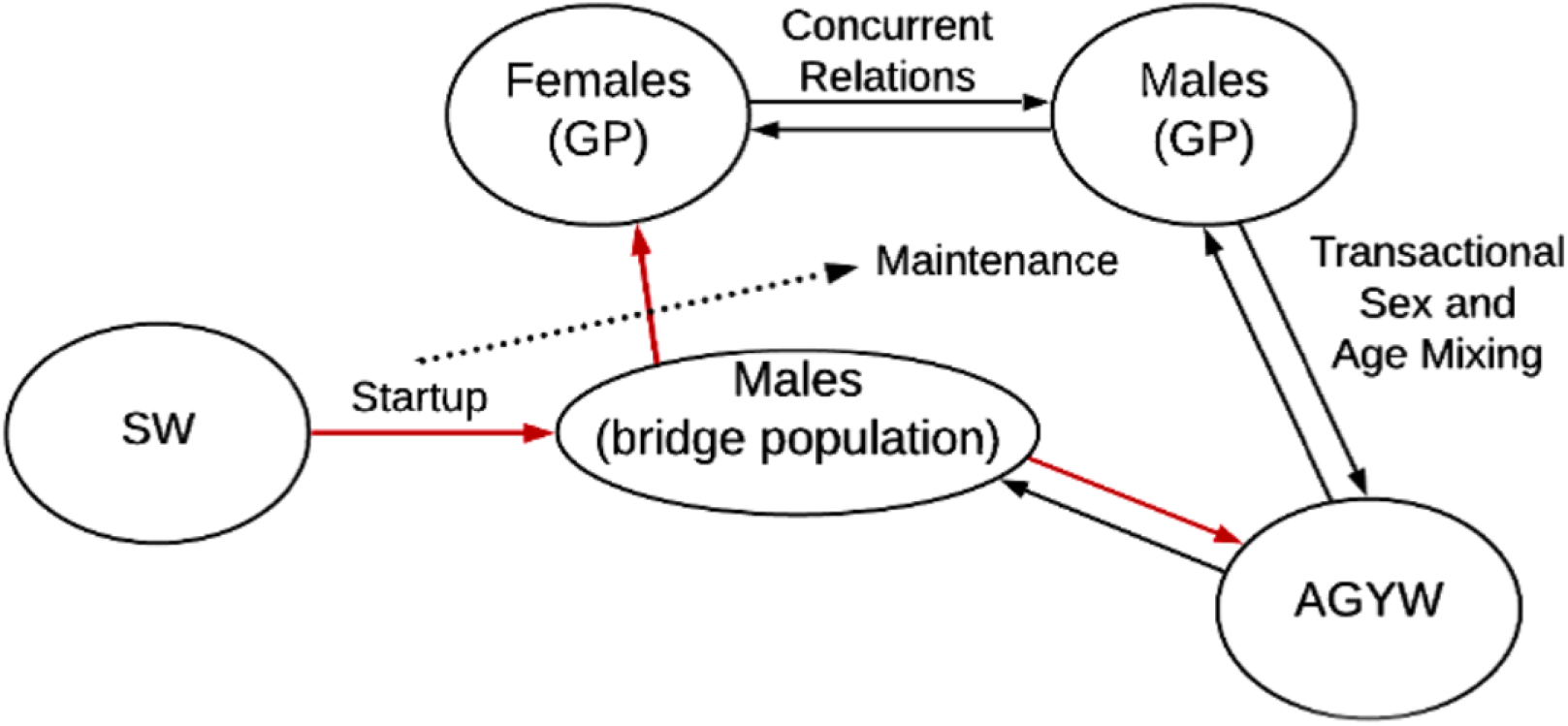
Conceptual model of HIV progression from concentrated epidemic to generalized and self-maintained epidemic in countries with the socio-behavioural characteristics of Madagascar^1^. 1. The start up of the epidemic is characterized by a long-lasting and steady increase of HIV prevalence among SWs. HIV infection spills over to GP (adult females, AGYW and indirectly other GP males) through bridge population (SW clients, see red arrows). The intensity of this initial spillover is enhanced as prevalence among SW increases in a positive feedback manner. Once a certain threshold in GP has been reached, prevalence/incidence in GP may be self-sustained and tend to increase through high risk intercourse between AGYW and older adults. This threshold may be reached sooner depending on the prevalence of age disparate relationships, concurrency and inconsistent use of condoms. Transactional sex may be the main mediator of such risk factors.

However, the generalization of the HIV epidemic is far from a homogeneous process across the 11 studied cities, and it looks more like a patchwork of micro-epidemics leading to different regional and subregional stages (Fig 2). In spite of this heterogeneity, if our projections are correct, future HIV prevalence in Madagascar for next decade (2030s) may be similar to other HIV/AIDS highly hit countries from the Southern African region between 9-24%, unless a sustained action is taken.

Our model is based on the key recognition of commercial sex as the main driver of HIV infection leading to a generalized epidemic in Sub-Saharan Africa [21]. However, we expanded the classical concept of sex workers, encompassing the consideration of transactional sex (occasional sexual intercourse in exchange of any material or non-material benefit other than money). Transactional sex has been underscored as a risky factor for HIV expansion [22], and it is especially prevalent among AGYW and well documented in Madagascar [2, 23]. A disproportionate fraction of sexual encounters between males and younger females (intercourse mixing age heterogeneity, factored by the parameter fW and f0, see Table SB1 and SB2, and also Fig SE7 in SI), has been early noticed as a risk factor of HIV acquisition for AGYW on itself, and through associated risky behaviors, such as concurrency and inconsistent use of condoms [24–26]. Thereby, AGYW is now considered as a truly key population in most Sub-Saharan countries [5], with six or sevenfold higher risk of HIV acquisition compared to males of the same age [26, 27]. In Madagascar, all of these factors and behaviors have been reported as highly prevalent [2]. Of note, the widespread presence of sex working and transactional sex is strongly influenced by underlying socio-economic factors and, since they are both driven by financial insecurity and poverty [26] and structural factors such violence and criminalization [28]. Consequently, our model may underestimate the speed of transition towards generalized epidemics if a major economic crisis occurs, as we expect as consequence of the current Covid-19 epidemic or climate change impacts in form of droughts and secondary food insecurity, as it has been recently experienced in Madagascar [29], which may in turn hamper the HIV response [30]. Indeed, it has been underlined the disproportionate susceptibility of women to these factors [31]. For this reason, it is striking to observe that future projections of HIV/AIDS epidemics focus almost entirely on sustained provision of anti-retroviral drugs and HIV-services [32, 33] without considering the impact of the Covid-19 crisis on the economy and other cofactors.

In agreement with previous work [34, 35], our model invariably predicts that the sex workers population is the first to take off in the trajectory towards disease establishment (Fig 5a). At that point, sex workers HIV prevalence may be the startup of the HIV introduction into the general population through bridge populations (clients), followed by a further expansion of HIV infection mediated by transactional sex mainly practiced by AGYW and older males. Therefore, the intercourse mixing-age model and risky behaviors associated to transactional sex may play a crucial role in the spread and maintenance of the epidemics once HIV prevalence has reached a certain threshold among general population (see Fig 6). At a certain stage, the HIV epidemics evolve independently from the number of HIV positive sex workers [34]. The inclusion of different infectiousness levels (see Fig 1) through the stages of the natural history of a HIV infection allows to explore their interaction with behavioral factors which are particularly associated to the described pattern of sexual encounters [36], and circumcision. Here, we strongly suggest that acute infections would play an important role in the epidemics transition. Accordingly, previous authors pointed out the disproportionate contribution of acute infections at the early stages of disease expansion [37]. Although most previous HIV compartmental models of this kind [38, 39] do not consider these critical interactions, previous work considering some of them leads to similar conclusions [35].

Main parameters of HIV transmission established by previous observational studies lie within the bounds of our model estimations. Scarce HIV cases has been reported in Madagascar at least since 1989 [40]. Therefore, we calculated R0 values in 2000, when prevalence was still thought to be very low. These values depend on most model parameters, and vary between about 3 and 9 across the different cities (Fig 2) in good agreement with observational estimations [41]. However, these estimations may be a little inflated because they are calculated under the assumption of random sexual contacts between the different groups [42]. Notwithstanding, these values are particularly useful for relative comparisons, and to elucidate which model parameters have the highest influence on disease propagation just after HIV introduction. They represent highest bounds, as it is known that in general contact heterogeneity, and non-random sexual mixing (for instance, sequential monogamy), commonly slows down disease propagation [43]. In summary, from our case-study, we suggest that factors influencing the transition towards a generalized HIV epidemics result from a complex interplay between geographic isolation, which may be enhanced by political issues, slowing down the introduction of HIV, cultural practices, such as circumcision, but as well the societal consideration of women and gender-based practices that may protect young women from risky behaviors (i.e. early marriage, matriarchal structures), biological (i.e. the key role of acute infections during the transitional phase), and socio-economic disruptions. The transition from rural to urban societies, with the interaction between all these elements, may exacerbate these underlying factors (erosion of traditional practices, increasing population mobility and lack of social protection towards AGYW).

Our study is limited first by the scarce information available on the distribution of women in the different key groups (Sex-workers and transactional sex, and non-sex-workers or -transactional sex). Specific hypotheses controlling this distribution are implicitly assumed in the set of α and σ’s parameters (see Fig 1 and Tables SA1, SB1 and SB2 in SI). Second, there is no direct data on the age-disparity of sexual relationships in Madagascar, and finally, the paucity of epidemiological time-series data, as pointed out previously [2]. These information gaps need to be filled to obtain more robust projections. Furthermore, we do not considered other key populations (men who have sex with men and injected drug users), which have been largely neglected [44], but their role in the generalization of epidemics should not be discarded. In fact, the role of injected drug users has been particularly underscored in Sub-Saharan Africa [45], and this represents an increasing population in Madagascar [2]. The bridge between these key population and the general population through bisexual intercourses and sex working, should be further explored. Finally, future models of this kind should also take into account mobility between localities as pointed out recently [46, 47]. At this stage, our model does not consider the movement of populations at risk (specifically sex workers) between cities following tourist seasons, as it has been reported in Mahajanga and Nosy-be (the two cities with higher HIV prevalence among sex workers) [2, 23]. These dynamics may foster even more diffusion of HIV to other parts of the country. Our projections should be considered a call for action given the scant attention that HIV in Madagascar has received, and the Public Health crisis that potentially could unfold, but could still be avoided. Furthermore, the lack of mobility between cities and a possible underestimation of the amount of AGYW practicing transactional sex could make our model projections still conservative. Even admitting that our model is based on simplified assumptions and limited data, the case-study of Madagascar can help to understand how has occurred (or may occur) the transition from concentrated to generalized epidemics in different settings. In this sense, considering our projections, it is worrisome to see that Madagascar shows some of the poorest HIV response indicators in the world (only 14% of persons living with HIV are estimated to be under follow-up and this data was not considered in our modeling estimates) [48]. Given the plausibility of the worst case scenario outlined in this article, it is important to set up observational studies on HIV prevalence/incidence and risky factors that would help to draft more accurate predictions, alongside the implementation of robust preventive measures focused on monitoring and identifying hot-spots and vulnerable key populations (sex workers and AGYW).

## Conclusion

The UNAIDS Fast Track Strategy aimed to end HIV epidemic by 2030. However, in spite of the outstanding progress experienced, like the sustained decline on AIDS-related deaths, the disease is far from being under control. Setbacks could still happen, including crisis with major socio-economical or management care disruptions due to the Covid-19 epidemic, climate change or other natural disasters. Disentangling the interplay between the different factors driving HIV expansion requires a dynamic approach. By developing a new data-based transmission dynamic model, here we report that Madagascar could be undergoing a silent, non-linear transition from a low-prevalence, concentrated to a well-established generalized epidemic. Unless a sustained response is implemented, we foresee that this country would reach HIV prevalence similar to high endemic countries by 2033. Not only in Madagascar, but also in other settings, our work can help to understand the context-dependent interactions underlying these rapid transitions and raise alarm bells before it is too late to avoid them.

## Data Availability

We do not have legal or ethical restrictions to make data fully available

https://doi.org/10.5061/dryad.3ffbg79mn

## Supporting Information Appendix (SI)

A PDF file includes detailed supporting information about the dynamical ODE system corresponding to the model (Fig 1), the R0 calculation, the strategy to conduct parameter estimation, model projections, and data sources. Baseline and intermediate data has been made fully accessible [14] and we included as supplementary material a txt file with instructions to disclose and analyze it independently.

## Author contributions

Both authors contributed equally to conceptualization, investigation, methodology, project administration, funding acquisition and resources, visualization, data, writing original draft, and reviewing and editing the final manuscript. XV contributed to data acquisition, and underlying data verification. DA contributed to software, formal analysis, model fitting and validation, and the preparation of the supporting information file. All authors have read and agreed to the submitted version of the manuscript.

## Author declaration

The authors declare no conflict of interest.

## Ethical statement

The study used aggregated data from previously published workds (see reference list) and did not included any sampling procedure with individual level information or the use of biological or personal data which would need the consent from individual participants. This study is part of a larger HIV initiative in Madagascar which had the clearance of the Ethics committee of the Ministry of Public health of Madagascar (number 088/MSANP/CERBM).

## Acknowledgements

This work was funded by the Spanish Ministerio de Ciencia, Innovacion y Universidades and the European Regional Development Fund (ERDF),under the project CRISIS (PGC2018-096577-B-I00 to D.A.). The field work, data acquisition and data revision of XV was supported by the non-governmental organization (NGO) Medecins du Monde (France) and was funded by Expertise France (EF) (Initiative 5%, ref. num. 16SANIN208). We also acknowledge the Theoretical and Computational Ecology group at the Center for Advanced Studies of Blanes (CEAB-CSIC) for providing a nice and supportive working environment. We are also very thankful to Armun Liaghat at the University of Chicago, where the initial ideas of this work were developed during a Tinker Visiting Professorship stay (to DA), Molebogeng Rangaka, from the University College of London and Laurence Baril for their observations, corrections, careful proofreading and useful comments.

## Notes

### Competing Interest Statement

The authors have declared no competing interest.

## References

1. Brown T, Peerapatanapokin W. Evolving HIV epidemics: the urgent need to refocus on populations with risk. Curr Opin HIV AIDS. 2019 Sep;14(5):337–353. doi: 10.1097/COH.0000000000000571.

2. Raberahona M, Monge F, Andrianiaina RH, Randria MJD, Ratefiharimanana A, Rakatoarivelo RA, Randrianary L, Randriamilahatra E, Rakotobe L, Mattern C, Andriananja V, Rajaonarison H, Randrianarisoa M, Rakotomanana E, Pourette D, Andriamahenina HZ, Dezé C, Boukli N, Baril L, Vallès X. Is Madagascar at the edge of a generalised HIV epidemic? Situational analysis. Sex Transm Infect. 2021 Feb;97(1):27–32. doi: 10.1136/sextrans-2019-054254.

3. Behets F, Andriamiadana J, Rasamilalao D, Ratsimbazafy N, Randrianasolo D, Dallabetta G, Cohen M. Sexually transmitted infections and associated socio-demographic and behavioural factors in women seeking primary care suggest Madagascar’s vulnerability to rapid HIV spread. Trop Med Int Health. 2001 Mar;6(3):202–11. doi: 10.1046/j.1365-3156.2001.00690.x. PMID: 11299037.

4. Anderson RM and May RM. Infectious Diseases of Humans. Dynamics and Control. Oxford University Press, Oxford, 1991.

5. Dellar RC, Dlamini S, Karim QA. Adolescent girls and young women: key populations for HIV epidemic control. J Int AIDS Soc. 2015 Feb 26;18(2 Suppl 1):19408. doi: 10.7448/IAS.18.2.19408.

6. Fiebig EW, Wright DJ, Rawal BD, Garrett PE, Schumacher RT, Peddada L, Heldebrant C, Smith R, Conrad A, Kleinman SH, Busch MP. Dynamics of HIV viremia and antibody seroconversion in plasma donors: implications for diagnosis and staging of primary HIV infection. AIDS. 2003 Sep 5;17(13):1871–9. doi: 10.1097/00002030-200309050-00005.

7. Hollingsworth TD, Anderson RM, Fraser C. HIV-1 transmission, by stage of infection. J Infect Dis. 2008 Sep 1;198(5):687–93. doi: 10.1086/590501.

8. Pinkerton SD. Probability of HIV transmission during acute infection in Rakai, Uganda. AIDS Behav. 2008 Sep;12(5):677–84. doi: 10.1007/s10461-007-9329-1.

9. Bacchetti P, Moss AR. Incubation period of AIDS in San Francisco. Nature. 1989 Mar 16;338(6212):251–3. doi: 10.1038/338251a0.

10. CIA. Central Intelligence Agency. https://www.cia.gov/library/publications/the-world-factbook/fields/2018.html, 2018. Accessed: September 2021

11. Instat. Institute de Statistique de Madagascar. https://www.instat.mg, 2018. Accessed: September 2021.

12. WHO. World HealthOrganization. http://apps.who.int/gho/data/node.home, 2018. Accessed: September 2021.

13. Worldometers. Worldometers. https://www.worldometers.info/world-population/madagascar-population/, 2018. Accessed: September 2021.

14. Alonso D, Valles X (2022). Supporting data for “A potential transition from a concentrated to a generalised HIV epidemic: the case of Madagascar”.Dryad, Dataset. https://doi.org/10.5061/dryad.3ffbg79mn

15. Alsallaq RA, Cash B, Weiss HA, Longini IM Jr, Omer SB, Wawer MJ, Gray RH, Abu-Raddad LJ. Quantitative assessment of the role of male circumcision in HIV epidemiology at the population level. Epidemics. 2009 Sep;1(3):139–52. doi: 10.1016/j.epidem.2009.08.001.

16. Sharma SC, Raison N, Khan S, Shabbir M, Dasgupta P, Ahmed K. Male circumcision for the prevention of human immunodeficiency virus (HIV) acquisition: a meta-analysis. BJU Int. 2018 Apr;121(4):515–526. doi: 10.1111/bju.14102.

17. Lei JH, Liu LR, Wei Q, Yan SB, Yang L, Song TR, Yuan HC, Lv X, Han P. Circumcision Status and Risk of HIV Acquisition during Heterosexual Intercourse for Both Males and Females: A Meta-Analysis. PLoS One. 2015 May 5;10(5):e0125436. doi: 10.1371/journal.pone.0125436.

18. Alonso D, Dobson A, Pascual M. Critical transitions in malaria transmission models are consistently generated by superinfection. Philos Trans R Soc Lond B Biol Sci. 2019 Jun 24;374(1775):20180275. doi: 10.1098/rstb.2018.0275.

19. Comité National de Lutte contre le VIH/Sida. Resultats de l’Enquête de surveillance Biologique du VIH/SIDA et de la Syphilis. Ministère de la Santé du Planning Familial et de la Protection Sociale, Madagascar. 2005. Available at https://www.scribd.com/document/48559097

20. Comité National de Lutte contre le VIH/Sida. Resultats de l’Enquête de surveillance Biologique du VIH/SIDA et de la Syphilis. Ministère de la Santé du Planning Familial et de la Protection Sociale, Madagascar. 2007. Available at https://www.scribd.com/document/48559097

21. Talbott JR. Size matters: the number of prostitutes and the global HIV/AIDS pandemic. PLoS One. 2007 Jun 20;2(6):e543. doi: 10.1371/journal.pone.0000543.

22. Wamoyi J, Stobeanau K, Bobrova N, Abramsky T, Watts C. Transactional sex and risk for HIV infection in sub-Saharan Africa: a systematic review and meta-analysis. J Int AIDS Soc. 2016 Nov 2;19(1):20992. doi: 10.7448/IAS.19.1.20992.

23. Freedman JJ, Rakotoarindrasata M. Remise en cause des frontières supposées entre travail du sexe et sexe transactionnel à Madagascar - le cas de Nosy-be. Technical report, Institut du Genre en Géopolitique, Paris, 2020. Available at https://igg-geo.org/?p=2434.

24. Evans M, Maughan-Brown B, Zungu N, George G. HIV Prevalence and ART Use Among Men in Partnerships with 15-29 Year Old Women in South Africa: HIV Risk Implications for Young Women in Age-Disparate Partnerships. AIDS Behav. 2017 Aug;21(8):2533–2542. doi: 10.1007/s10461-017-1741-6.

25. Maughan-Brown B, Evans M, George G. Sexual Behaviour of Men and Women within Age-Disparate Partnerships in South Africa: Implications for Young Women’s HIV Risk. PLoS One. 2016 Aug 15;11(8):e0159162. doi: 10.1371/journal.pone.0159162.

26. Leclerc-Madlala S. Age-disparate and intergenerational sex in southern Africa: the dynamics of hypervulnerability. AIDS. 2008 Dec;22 Suppl 4:S17–25. doi: 10.1097/01.aids.0000341774.86500.53.

27. Birdthistle I, Tanton C, Tomita A, de Graaf K, Schaffnit SB, Tanser F, Slaymaker E. Recent levels and trends in HIV incidence rates among adolescent girls and young women in ten high-prevalence African countries: a systematic review and meta-analysis. Lancet Glob Health. 2019 Nov;7(11):e1521–e1540. doi: 10.1016/S2214-109X(19)30410-3.

28. Shannon K, Strathdee SA, Goldenberg SM, Duff P, Mwangi P, Rusakova M, Reza-Paul S, Lau J, Deering K, Pickles MR, Boily MC. Global epidemiology of HIV among female sex workers: influence of structural determinants. Lancet. 2015 Jan 3;385(9962):55–71. doi: 10.1016/S0140-6736(14)60931-4.

29. Makoni M. Southern Madagascar faces “shocking” lack of food. Lancet. 2021 Jun 12;397(10291):2239. doi: 10.1016/S0140-6736(21)01296-4.

30. Orievulu KS, Ayeb-Karlsson S, Ngema S, Baisley K, Tanser F, Ngwenya N, Seeley J, Hanekom W, Herbst K, Kniveton D, Iwuji CC. Exploring linkages between drought and HIV treatment adherence in Africa: a systematic review. Lancet Planet Health. 2022 Apr;6(4):e359–e370. doi: 10.1016/S2542-5196(22)00016-X.

31. Austin KF, Noble MD, Berndt VK. Drying Climates and Gendered Suffering: Links Between Drought, Food Insecurity, and Women’s HIV in Less-Developed Countries. Soc Indic Res. 2021;154(1):313–334. doi: 10.1007/s11205-020-02562-x.

32. Jewell BL, Mudimu E, Stover J, Ten Brink D, Phillips AN, Smith JA, Martin-Hughes R, Teng Y, Glaubius R, Mahiane SG, Bansi-Matharu L, Taramusi I, Chagoma N, Morrison M, Doherty M, Marsh K, Bershteyn A, Hallett TB, Kelly SL; HIV Modelling Consortium. Potential effects of disruption to HIV programmes in sub-Saharan Africa caused by COVID-19: results from multiple mathematical models. Lancet HIV. 2020 Sep;7(9):e629–e640. doi: 10.1016/S2352-3018(20)30211-3.

33. The Lancet. Maintaining the HIV response in a world shaped by COVID-19. Lancet. 2020 Nov 28;396(10264):1703. doi: 10.1016/S0140-6736(20)32526-5.

34. Bershteyn A, Klein DJ, Eckhoff PA. Age-dependent partnering and the HIV transmission chain: a microsimulation analysis. J R Soc Interface. 2013 Aug 28;10(88):20130613. doi: 10.1098/rsif.2013.0613. PMID: 23985734; PMCID: PMC3785829.

35. Vickerman P, Foss AM, Pickles M, Deering K, Verma S, Eric Demers, Lowndes CM, Moses S, Alary M, Boily MC. To what extent is the HIV epidemic in southern India driven by commercial sex? A modelling analysis. AIDS. 2010 Oct 23;24(16):2563–72. doi: 10.1097/QAD.0b013e32833e8663.

36. Mah TL, Shelton JD. Concurrency revisited: increasing and compelling epidemiological evidence. J Int AIDS Soc. 2011 Jun 20;14:33. doi: 10.1186/1758-2652-14-33. PMID: 21689437; PMCID: PMC3133533.

37. Eaton JW, Hallett TB, Garnett GP. Concurrent sexual partnerships and primary HIV infection: a critical interaction. AIDS Behav. 2011 May;15(4):687–92. doi: 10.1007/s10461-010-9787-8.

38. Silva CJ, Torres DFM. A SICA compartmental model in epidemiology with application to HIV/AIDS in Cape Verde. Ecological Complexity. 2017; 30:70–75. doi: 10.1016/j.ecocom.2016.12.001.

39. Omondi EO, Mbogo RW, Luboobi LS. A mathematical modelling study of HIV infection in two heterosexual age groups in Kenya. Infect Dis Model. 2019 Apr 21;4:83–98. doi: 10.1016/j.idm.2019.04.003.

40. Andriamahenina R, Ravelojaona B, Rarivoharilala E, Ravaoarimalala C, Andriamiadana J, Andriamahefazafy B, May JF, Behets F, Rasamindrakotroka A. Le SIDA à Madagascar. I. Epidémiologie, projections, impact socio-économique, interventions [AIDS in Madagascar. I. Epidemiology, projections, socioeconomic impact, interventions]. Bull Soc Pathol Exot. 1998;91(1):68–70. French. PMID: 9559168.

41. Nsubuga RN, White RG, Mayanja BN, Shafer LA. Estimation of the HIV basic reproduction number in rural south west Uganda: 1991-2008. PLoS One. 2014 Jan 3;9(1):e83778. doi: 10.1371/journal.pone.0083778.

42. Diekmann O, Heesterbeek JA, Roberts MG. The construction of next-generation matrices for compartmental epidemic models. J R Soc Interface. 2010 Jun 6;7(47):873–85. doi: 10.1098/rsif.2009.0386.

43. Britton T, Ball F, Trapman P. A mathematical model reveals the influence of population heterogeneity on herd immunity to SARS-CoV-2. Science. 2020 Aug 14;369(6505):846–849. doi: 10.1126/science.abc6810.

44. Jin H, Restar A, Beyrer C. Overview of the epidemiological conditions of HIV among key populations in Africa. J Int AIDS Soc. 2021 Jul;24 Suppl 3(Suppl 3):e25716. doi: 10.1002/jia2.25716.

45. Asher AK, Hahn JA, Couture MC, Maher K, Page K. people who inject drugs, HIV risk, and HIV testing uptake in sub-Saharan Africa. J Assoc Nurses AIDS Care. 2013 Nov-Dec;24(6):e35–44. doi: 10.1016/j.jana.2012.09.003.

46. Okano JT, Sharp K, Valdano E, Palk L, Blower S. HIV transmission and source-sink dynamics in sub-Saharan Africa. Lancet HIV. 2020 Mar;7(3):e209–e214. doi: 10.1016/S2352-3018(19)30407-2. Epub 2020 Feb 14. PMID: 32066532; PMCID: PMC7259817.

47. Okano JT, Busang L, Seipone K, Valdano E, Blower S. The potential impact of country-level migration networks on HIV epidemics in sub-Saharan Africa: the case of Botswana. Lancet HIV. 2021 Nov 10:S2352-3018(21)00267-8. doi: 10.1016/S2352-3018(21)00267-8. Epub ahead of print. PMID: 34774183.

48. UNAIDS; Country Fact Sheet Madagascar 2020. Available at https://www.unaids.org/en/regionscountries/countries/madagascar. Accessed on July 2022.

